# Effect of the COVID-19 pandemic in stroke code activations in the region of Madrid: a retrospective cohort study

**DOI:** 10.1101/2020.12.10.20246892

**Authors:** Nicolás Riera-López, Blanca Fuentes-Gimeno, Jorge Rodriguez-Pardo de Donlebun

## Abstract

Acute Stroke (AS) is the most common time-dependent disease attended in the Emergency Medicine Service (EMS) of Madrid (SUMMA 112). Community of Madrid has been one of the most affected regions in Spain by the coronavirus disease 2019 (COVID19) pandemic. A significant reduction in AS hospital admissions has been reported during the COVID-19 pandemic compared to the same period one year before. As international clinical practice guidelines support those patients with suspected acute stroke should be accessed via EMS, it is important to know whether the pandemic has jeopardized urgent pre-hospital stroke care, the first medical contact for most patients. We aimed to examine the impact of the COVID-19 in stroke codes (SC) in our EMS among three periods of time: the COVID-19 period, the same period the year before, and the 2019-2020 seasonal influenza period. Methods: We compared the SC frequency among the periods with high cumulative infection rate (above the median of the series) of the first wave of COVID-19, seasonal influenza, and also with the same period of the year before. Results: 1,130 SC were attended during the three periods. No significant reduction in SC was found during the COVID-19 pandemic. The reduction of hospital admissions might be attributable to patients attending the hospital by their means. The maximum SC workload seen during seasonal influenza has not been reached during the pandemic. We detected a non-significant deviation from the SC protocol, with a slight increase in hospitals’ transfers to hospitals without stroke units.

## INTRODUCTION

Acute Stroke (AS) is the most common time-dependent pathology attended in the Emergency Medicine Service (EMS) of Madrid (SUMMA 112). It is the first cause of mortality among women and the first cause of disability in adults.

Community of Madrid has been one of the most affected in Spain by the coronavirus disease 2019 (COVID-19) pandemic. Until May 31st, 68.830 cases and 8.691 fatalities have been registered, nearly one-third of the entire country’s total count.

Several studies reported an alarming decrease in hospital admissions for stroke [1-5]. But little information was available about the performance of pre-hospital EMS. International guidelines support patient access via EMS, following the stroke code protocol (SC) [6, 7]. This significantly reduces door-to-needle time and improves patient prognosis. A reduction of SC activation cases (usually the most severe stroke cases) would be a dangerous consequence of COVID-19.

EMS are used to dealing with other work overload situations such as the annual seasonal influenza (health system overload, similar severity risk factors). We hypothesized that the SC protocol has been quite strong despite the high incidence of COVID-19 during the first months of the pandemic. We aimed to study the frequency of SC compared with the previous year, and with the seasonal influenza period.

## METHODS

We conducted a retrospective observational cohort study including all consecutive acute stroke code patients attended by SUMMA 112, the leading Emergency Medical Service (EMS) in Madrid. We analyzed the daily frequency of SC and hospital destination considering three cohorts according to:

- November 24th, 2019, to February 14th, 2020, corresponding to the seasonal influenza period (incidence of infection above the median of the series).
- March 1st to May 8th, 2020, for the COVID-19 period (incidence of infection above the median of the series),
- March 1st to May 8th, 2019, for the PreCOVID-19 period (the same period the year before),

We selected the dates defining each period according to the official data on the daily cumulative incidence of infection above the median of the series of influenza (influenza sentinel system from the Institute of Health Carlos III) and COVID-19 (Spanish Health Ministry).

### Study data

Age and sex of the patients, daily SC frequency, and type of hospital destination (stroke-ready hospital, hospital with stroke unit, and hospital with stroke unit and endovascular facilities) from the SUMMA 112 prospective registry of stroke codes.

### Statistical analysis

We used RStudio for statistical analyses and Microsoft Excel for the graphs.

Three groups’ means of SC frequency have been compared using the ANOVA test with the Tukey-Kramer method for pairwise comparisons. The rest of the variables have been compared by ANOVA or Kruskal-Wallis tests as appropriate. Chi-square test has been used to compare categorical variables. The difference of means has been estimated with a 95% confidence interval, considering statistically significant those with p<0,05.

This study was approved by the Ethics Committee of the Community of Madrid. As a retrospective study, patient consent was waived.

## RESULTS

SUMMA 112 attends an average of 150 SC (95% CI: 133-167) per month (figure 1). We attended 1,130 SC during the three periods.

**Figure 1:**
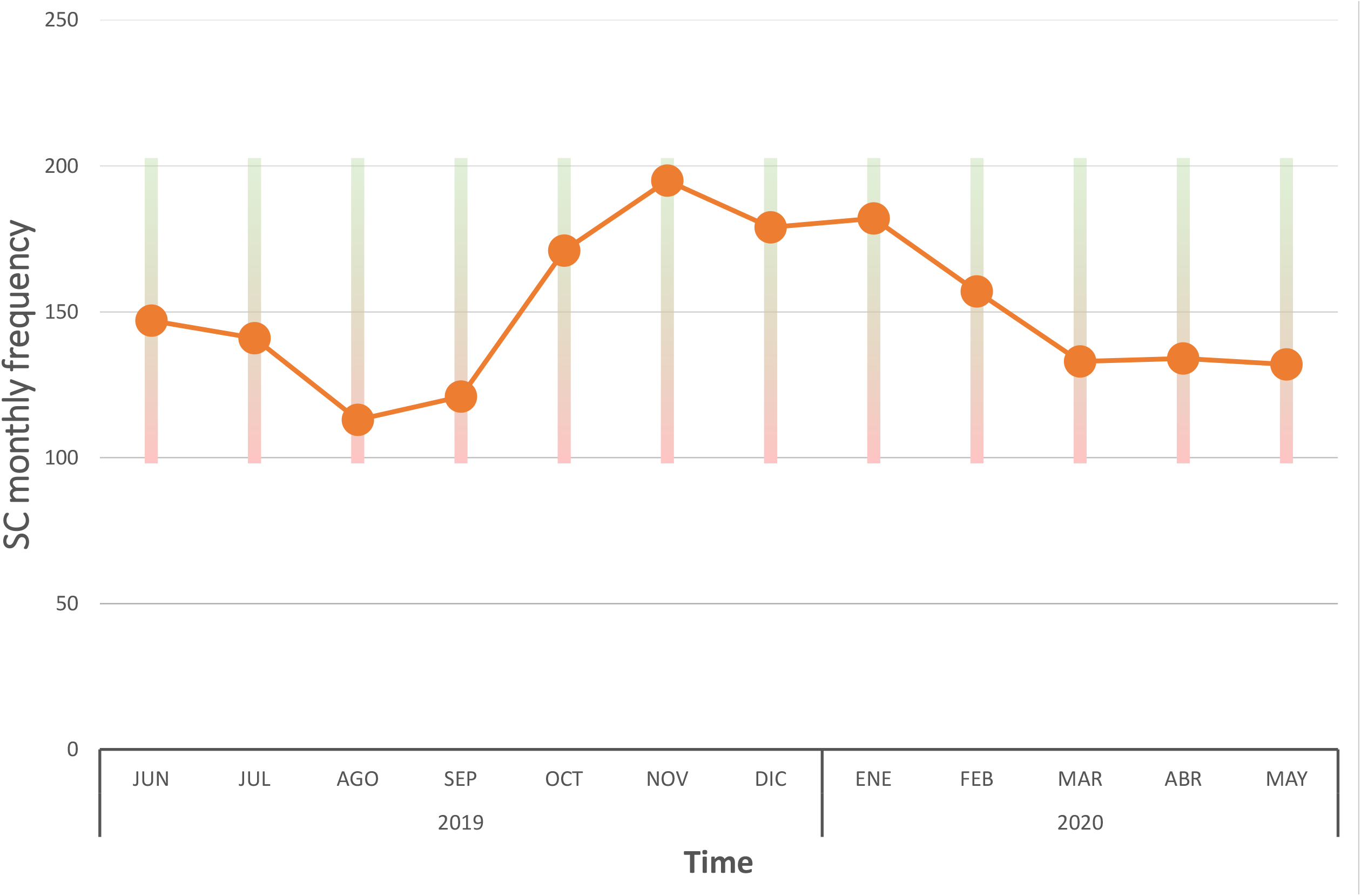
Monthly frequency of Stroke Codes in SUMMA 112 (shadowed area 95% CI).

During the period of COVID-19, a non-significant daily reduction of 13% SC was observed compared with the pre-COVID-19 period (difference of -0.68 daily cases, 95% CI: 0.15 to -1.51) and a statistically significant reduction of 24% compared with the seasonal influenza period (−1.41, 95% CI: -0.62 to -2.21) (figure 2).

**Figure 2:**
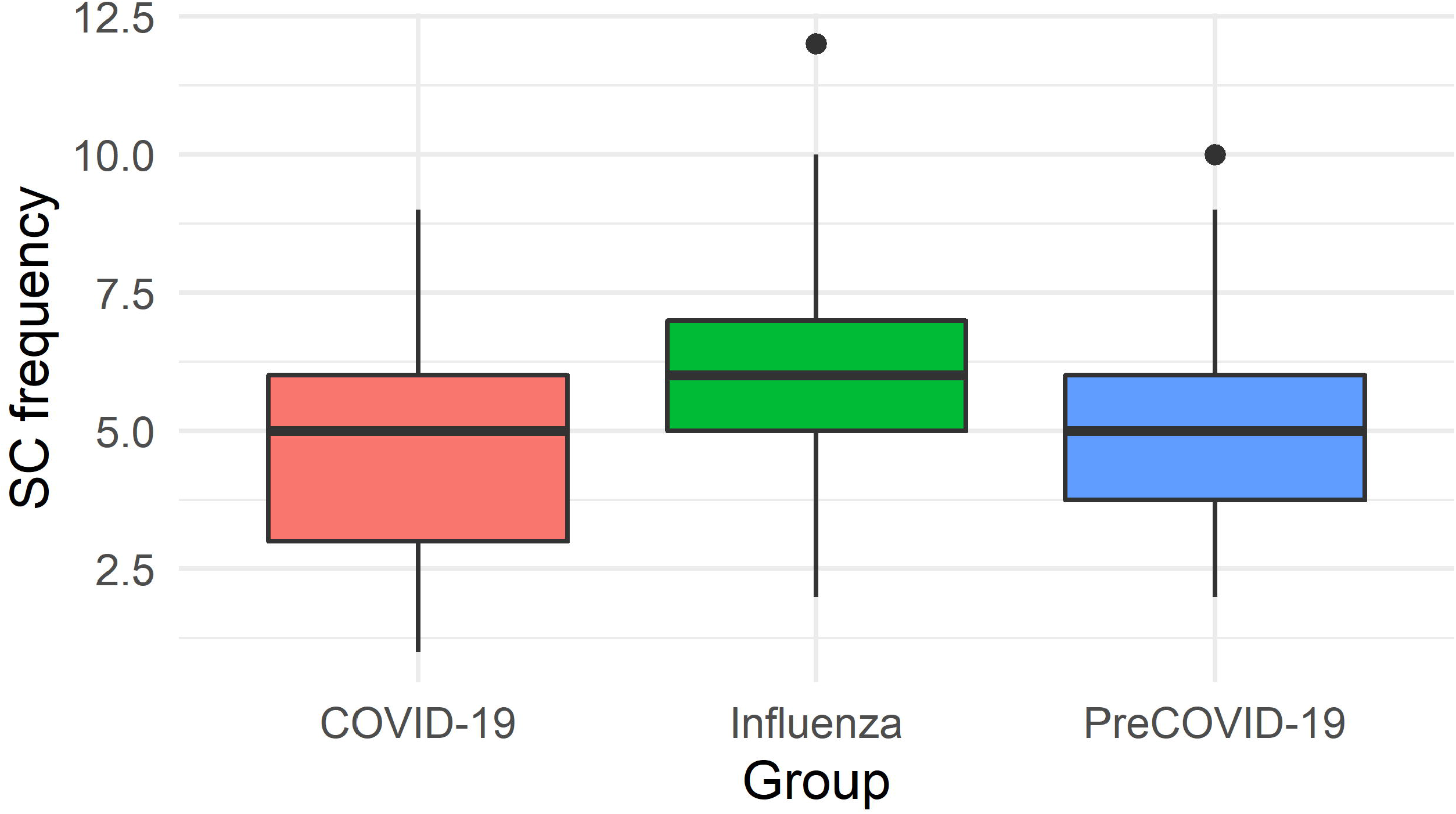
Daily frequency of stroke codes in each of the periods.

A significant younger age of the COVID-19 group (95% CI: 67.6-71) compared with the other two groups (pre-COVID-19, 95%CI: 71.2-74; influenza, 95% CI: 71.8-74.3) was found. Also, a lower proportion of women were attended with suspected stroke (COVID-19: 43.9%, 95% CI: 38.2%-49.7%; pre-COVID-19 51.9%, 95% CI: 46.4% -57.2%; influenza 53.3%, 95% CI: 48.8%-57.8%) (table 1).

**Table 1.**
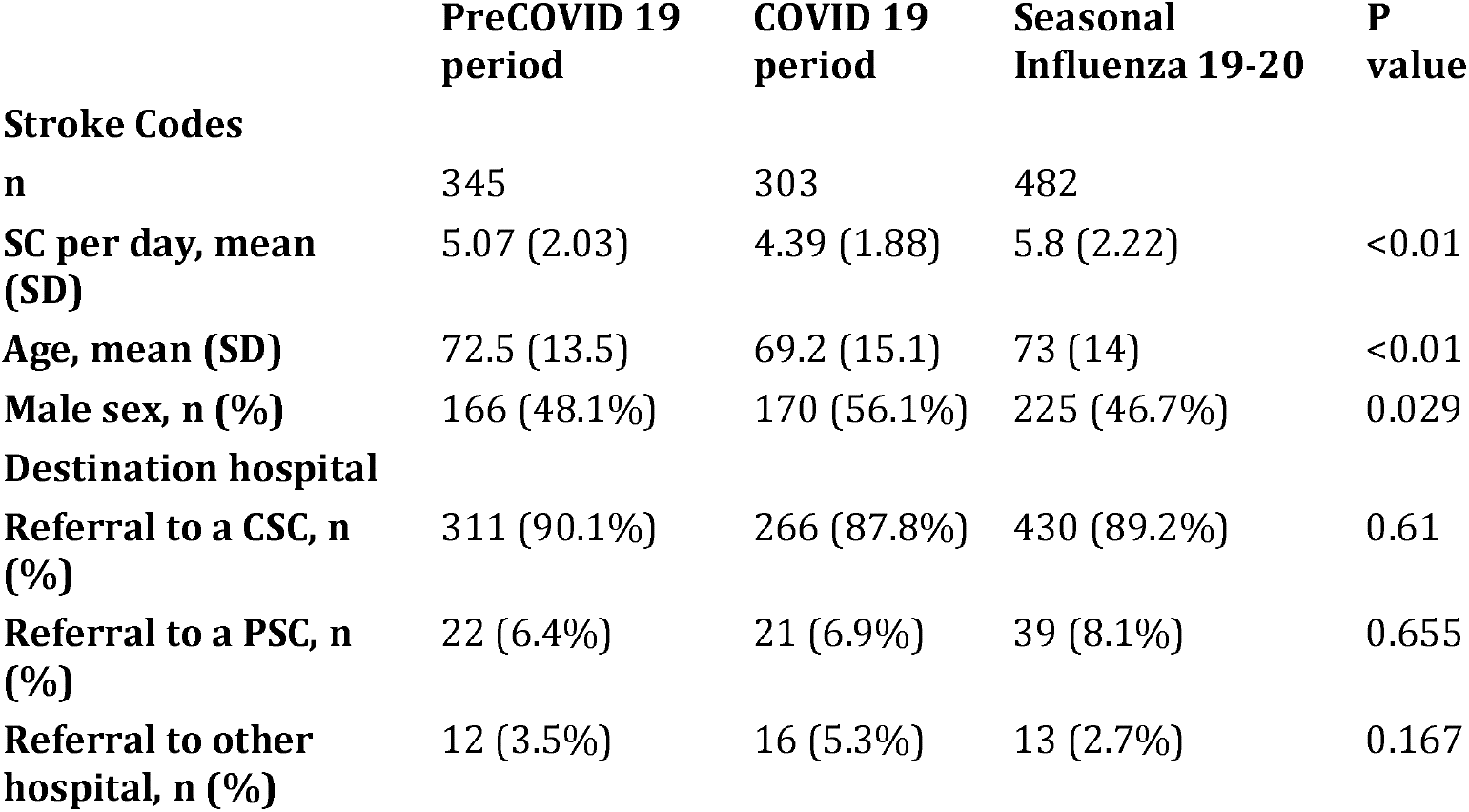

|  | **PreCOVID 19**<br>**period** | **COVID 19**<br>**period** | **Seasonal**<br>**Influenza 19-20** | **P**<br>**value** |
| --- | --- | --- | --- | --- |
| **Stroke Codes** |  |  |  |  |
| **n** | 345 | 303 | 482 |  |
| **SC per day, mean**<br>**(SD)** | 5.07 (2.03) | 4.39 (1.88) | 5.8 (2.22) | <0.01 |
| **Age, mean (SD)** | 72.5 (13.5) | 69.2 (15.1) | 73 (14) | <0.01 |
| **Male sex, n (%)** | 166 (48.1%) | 170 (56.1%) | 225 (46.7%) | 0.029 |
| **Destination hospital** |  |  |  |  |
| **Referral to a CSC, n**<br>**(%)** | 311 (90.1%) | 266 (87.8%) | 430 (89.2%) | 0.61 |
| **Referral to a PSC, n**<br>**(%)** | 22 (6.4%) | 21 (6.9%) | 39 (8.1%) | 0.655 |
| **Referral to other**<br>**hospital, n (%)** | 12 (3.5%) | 16 (5.3%) | 13 (2.7%) | 0.167 |

Finally, there was also a non-significant reduction in transfer rates to stroke centers (87% vs. 90%, p=0.61) at the cost of increasing transfers to hospitals not suitable for SC (5% vs. 3%, p=0.17).

## DISCUSSION

Stroke admissions in hospitals are a sum of those transferred by the EMS and those arriving by their means. We did not detect a significant decrease in SC cases during the COVID-19 period (figure 2). So, the main reason for the reported reduction of stroke admissions might be in the latter group. It is congruent with the reported reduction in transient ischemic attack and mild strokes [8]. Patients with mild or temporary symptoms may have been more concerned about being infected in the hospital or having delayed their evaluation. Efforts should be made in awareness campaigns, in the message to call 112 whenever the patient identifies a sign of stroke.

An increase in SC has also not been detected, despite the evidence of thrombotic events associated with SARS-CoV-2 infection [9]. One reason pointed out by other authors is the saturation of the EMS resources, including emergency hotlines that might have blocked potential patients with stroke symptoms from being attended [1].

Stroke patients were younger in the COVID-19 period. Older people may have tried to avoid attending the emergency department and strictly followed the messages of staying at home [1-2].

We want to highlight the stability of the SC protocol that has succeeded under those extraordinary circumstances, although we detected a non-significant deviation with a slight increase in transfers to hospitals without stroke units.

To our knowledge, this is one of the first studies to analyze the behavior of pre-hospital EMS in a time-dependent pathology as prevalent as AS. The major limitation of this study is that we focus it on calls to 112, and the information on other AS patients is lacking. We de did not find evidence of a significant reduction of SC during the COVID-19 pandemic compared with the same period of the year before. The number of SC was significantly lower during the COVID-19 period than the 2019-2020 seasonal influenza period, thus SC seems to operate correctly, even with a high level of health system saturation.

## Data Availability

Data available upon a reasonable request

## Acknowledgements

The authors thank the SUMMA-112 Coordination Center and the Community of Madrid Health Service (Servicio Madrileño de Salud) for making this possible. Special thanks to Isabel del Cura (Unidad de investigacion Gerencia Atencion Primaria Madrid - Fundación para la Investigación y la Innovación Biosanitaria en Atención Primaria de la Comunidad de Madrid, FIIBAP), for the manuscript review; Nuria Rodríguez Rodill (IT department of the Emergency Medical Service, SUMMA 112) for the data acquisition, and Balvinder Singh Powar for the manuscript translation.

